# SARS-CoV-2 transmission potential and rural-urban disease burden disparities across Alabama, Louisiana, and Mississippi, March 2020 — May 2021

**DOI:** 10.1101/2021.12.18.21268032

**Authors:** Sylvia K. Ofori, Chigozie A. Ogwara, Seoyon Kwon, Xinyi Hua, Kamryn M. Martin, Arshpreet Kaur Mallhi, Felix Twum, Gerardo Chowell, Isaac C.-H. Fung

**Affiliations:** Department of Biostatistics, Epidemiology and Environmental Health Sciences, Jiann-Ping Hsu College of Public Health, Georgia Southern University, Statesboro, GA 30458; Department of Health Sciences and Kinesiology, Waters College of Health Professions, Statesboro, GA 30458; The Dr. Lynn Cook Hartwig Public Health Program, The University of Southern Mississippi, Hattiesburg, MS 39406; Department of Population Health Sciences, School of Public Health, Georgia State University, Atlanta, GA 30303

**Keywords:** COVID-19, SARS-CoV-2, Reproduction number, Non-pharmaceutical interventions, Vaccine

## Abstract

**Purpose:** To quantify and compare SARS-CoV-2 transmission potential across Alabama, Louisiana, and Mississippi and selected counties with populations in the 50th, 75th, and 100th percentile.

**Methods:** To determine the time-varying reproduction number R_t_ of SARS-CoV-2, we applied the R package EpiEstim to the time series of daily incidence of confirmed cases. Median R_t_ percentage change when policies changed was determined. Linear regression was performed between log_10_-transformed cumulative incidence and log_10_-transformed population size at four time points.

**Results:** Stay-at-home orders, face mask mandates, and vaccinations were associated with the most significant reductions in SARS-CoV-2 transmission in the three southern states. R_t_ across the three states decreased significantly by 20% following stay-at-home orders. We observed varying degrees of reductions in R_t_ across states following other policies. Rural Alabama counties experienced higher per capita cumulative cases relative to urban ones as of June 17 and October 17, 2020. Meanwhile, Louisiana and Mississippi saw the disproportionate impact of SARS-CoV-2 in rural counties compared to urban ones throughout the study period.

**Conclusion:** State and county policies had an impact on local pandemic trajectories. The rural-urban disparities in case burden call for evidence-based approaches in tailoring health promotion interventions and vaccination campaigns to rural residents.

## Introduction

As of May 17, 2021, there were more than three million reported cases of coronavirus disease 2019 (COVID-19) in the United States (US), with over 30,000 new cases a day and almost 600,000 deaths.^1^ Evidence suggests disparities in COVID-19 burden across US census regions; the Southern region reported the second-highest cases with the most significant percentage increase during the early months of the pandemic.^2^ The southern US experienced surges in cases over the summer of 2020, partly driven by infection of younger adults,^3^ probably due to non-compliance to Centers for Disease Control and Prevention (CDC) guidelines.^4^ Despite the intensity of COVID-19 transmission in this region, compliance with social distancing measures was reported to be low.^5^

Alabama, Mississippi and Louisiana are three Southern Gulf states that have been heavily affected by the COVID-19 pandemic. These states are franked by Florida to the east and Texas to the west (see Supplementary Figures S1 and S2). Mississippi reported their first case on March 11, 2020,^6^ followed by Alabama on March 13, 2020,^7^ and finally Louisiana on March 14, 2020.^8^

To curb the pandemic, reaching the herd immunity threshold through vaccination is key.^9^ However, vaccine hesitancy presents a challenge in the South.^10^ The three Gulf states studied here had the lowest vaccination rates; <40% of the population had received at least one dose as of June 7, 2021.^11^

Rural-urban disparities in COVID-19 burden have been established in the literature.^12,13^ A study found that urban Louisianans had a significantly higher risk (Adjusted Relative Risk: 1.32, 95% CI, 1.22-1.43) of COVID-19 infection than rural Louisianans.^14^ In Mississippi, studies suggested that approximately half of the COVID-19-attributed hospitalizations were in rural areas as of April 25, 2020.^15,16^ The disparity was also true for Alabama.^17^ All three states had a higher percentage of residents living in rural areas than the national percentage (i.e., 14%): 23.16% for Alabama, 15.97% for Louisiana, and 53.17% for Mississippi, as of 2019.^18-21^ Research suggested that approaches to managing COVID-19 should account for rural-urban disparities including behavioral differences.^16,22^

Time-varying reproduction number (R_t_) is the average number of secondary cases generated by a typical infectious individual in the presence of public health interventions, behavioral changes, and increase in population immunity level. Hence, R_t_ changes over time throughout an epidemic. R_t_ estimation informs policymakers about how implemented policies and behavioral changes at the state and county level would impact COVID-19 transmission. When R_t_ >1, transmission is sustained, whereas when R_t_ <1, the epidemic will eventually die out.^23,24^

In this study, we explored the impact of different policies on SARS-CoV-2 transmission potential at the state level in Alabama, Louisiana, and Mississippi and evaluated rural-urban transmission differences using a representative set of counties with median, 75th, and 100th percentile population size.

## Methods

This study used retrospective data from the COVID-19 pandemic in Alabama, Louisiana, and Mississippi. The cumulative incidence data for each county in Alabama, Louisiana, and Mississippi were downloaded from the New York Times GitHub data repository up till May 17, 2021.^1^ The first COVID-19 cases were reported on March 13, 2020, in Alabama, March 9, 2020, in Louisiana, and March 11, 2020, in Mississippi. The daily number of new cases was obtained from the reported cumulative incidence by calculating the difference between consecutive cumulative case counts (Text S1). Executive orders of state administrations and timing for the implementation of policies were obtained from government and news sources.

### County Selection

Three counties were selected for each state based on population sizes and >10 daily new cases since ≤10 daily counts lead to unreliable R_t_ estimates.^25^ Counties at the median, 75th, and 100th percentiles as defined by the 2019 county-level population data from the US Census Bureau were selected:^26^ Chambers (median), Cullman (75^th^ percentile), and Jefferson (100^th^ percentile; County Seat: Birmingham) in Alabama; Evangeline (median), Iberia (75^th^ percentile), and East Baton Rouge (100^th^ percentile; Parish Seat: Baton Rouge) in Louisiana; and Leake (median), Marshall (75^th^ percentile), and Hinds (100^th^ percentile; County Seats: Raymond and Jackson) in Mississippi. Counties with population below the median were not analyzed here, as preliminary analysis found that the low case count in counties with small population size rendered the R_t_ estimates generated by the EpiEstim package very uncertain.

### Statistical analysis

R_t_ was estimated using the instantaneous reproduction number method.^23^ The EpiEstim package version 2.2-4 in R version 4.1.0 was used for the analysis.^27^ The serial interval distribution was parametrically defined (mean = 4.60 days; standard deviation = 5.55 days).^28^ The time series was shifted by nine days to approximate the onset of infection by assuming a mean incubation period of six days and a median testing delay of three days.^25,29-31^

Two sets of time window arrangements were used. First, the 7-day sliding window was used to minimize the fluctuations observed with smaller time steps by taking the average of R_t_ estimates over a week. Secondly, the non-overlapping time window method between which a bundle of interventions was implemented was used to estimate the average R_t_ over a given period.

To assess the extent of change in the R_t_ after policies were implemented, the percentage change was calculated for the non-overlapping time window R_t_ using the formula: 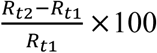. R_t2_ refers to the R_t_ estimate of the time window after a new policy was implemented and R_t1_ refers to the previous window. The sample_posterior_R(), *sample from the posterior R distribution*, function was used to sample 1000 estimates of R_t_ for each interval in the EpiEstim package and the associated 95% Credible interval (CrI) of the percentage change was calculated using bootstrapping. The percentage changes are presented in Figure 2 and Tables S1 through S4.

We explored the power-law relationship between the population size of counties and per capita cumulative case count using linear regression between the log_10_-transformed per capita cumulative case count and the log_10_-transformed population size. A negative slope indicates that counties with lower population size were associated with higher case burden while a positive slope means the opposite (Text S1).^32,33^ Time variability was assessed by regressing data at four time points (Date of report: June 17, 2020, October 17, 2020, February 17, 2021, and May 17, 2021).

### Ethics

The Georgia Southern University Institutional Review Board made a non-human subject determination for this project (H20364) under the G8 exemption category according to the Code of Federal Regulations Title 45 Part 46.

## Results

The daily number of new cases peaked twice in July and December 2020 in Alabama and Mississippi while the epidemic curve peaked thrice in April, July, and December 2020 in Louisiana (Figure 1). The cumulative case count per 10,000 population and the cumulative case count of each county of the three states are presented in maps (Figures S1 and S2). Detailed results can be found in Text S2.

**Figure 1:**
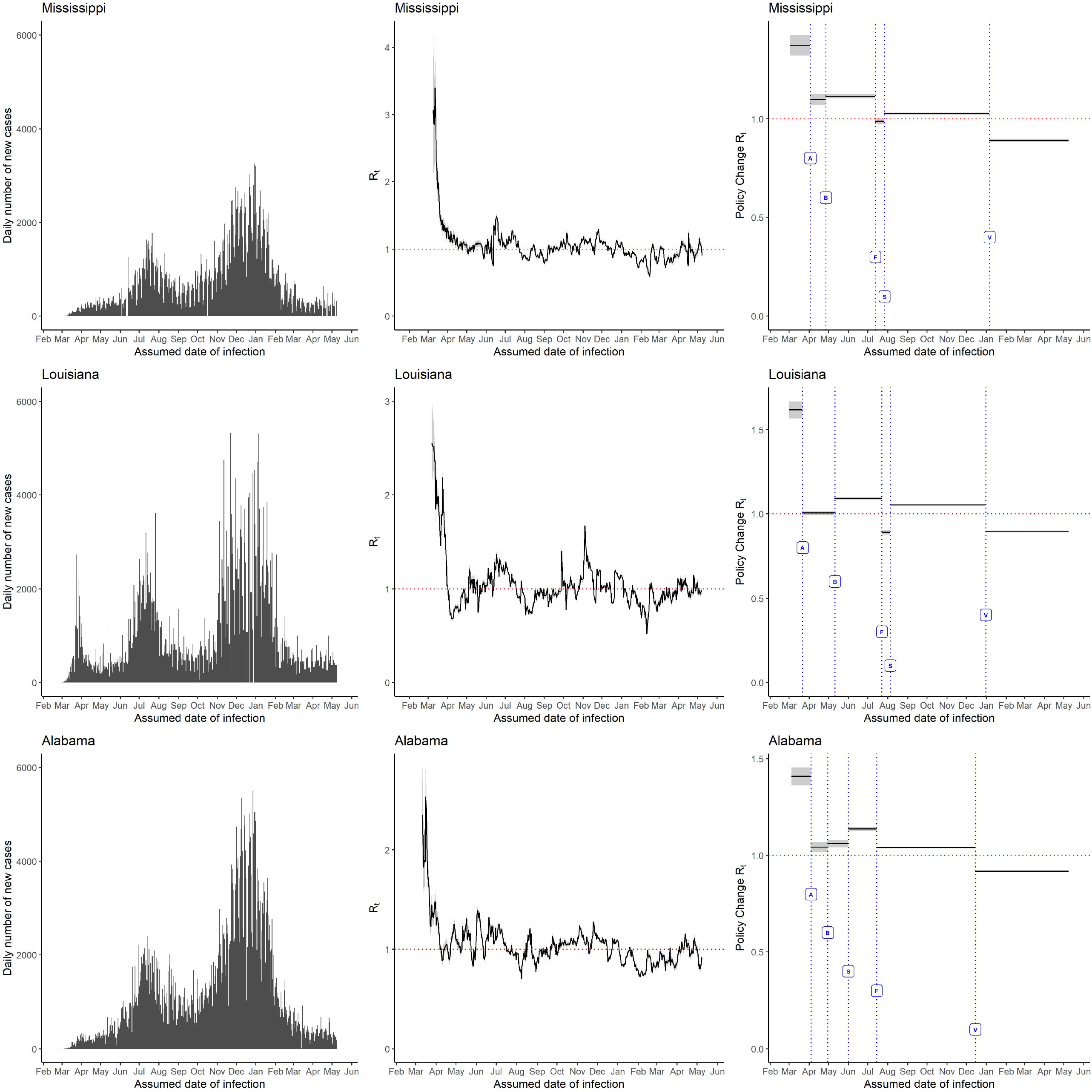
The daily number of new cases (left panel), 7-day sliding window Rt, (middle panel), and non-overlapping window Rt for policy change (right panel) for Alabama, Louisiana, and Mississippi. The government policies represented by the alphabets in the figure are: A= Stay at home order directive, B= Shelter in place/safer at home, S= School reopening, F= Face mask mandate and V= Rollout of vaccination began.

### 7-day sliding window R_t_ estimates at the state level

The 7-day sliding window R_t_ for all three states was above >1 in April 2020 but dropped to <1 between April and May for Louisiana before increasing again in late May. Alabama maintained a value >1 before dropping <1 in late May then fluctuated around 1 until December. Mississippi experienced similar fluctuations until December 2020. In all three states, the R_t_ was <1 between February and April 2021 but experienced a surge in May 2021 (Figure 1).

### 7-day sliding window R_t_ estimates at the county level

In Alabama, the R_t_ for Jefferson decreased to <1 in March 2020 and then fluctuated around 1 until May 2021; the R_t_ for Cullman and Chambers followed a similar trajectory. In Louisiana, all three selected parishes had peaks of R_t_ >3 in March, June, and November 2020; R_t_ decreased to <1 in April 2020 and generally fluctuated around 1 until May 2021. The selected counties in Mississippi followed a trend similar to the counties in Alabama and Louisiana (Figures S3, S4 and S5, Text S2).

### Policy impacts at the state level

The impact of policy changes on the transmission potential of SARS-CoV-2 as represented by the non-overlapping time window R_t_ are summarized in Figures 1, 2, and Table S1. Alabama, Louisiana, and Mississippi followed a similar trajectory in the changes in R_t_ as state orders were executed. The Stay-at-Home orders (represented as the letter A: enacted on April 4, 2020, in Alabama, on March 22, 2020, in Louisiana, and on April 3, 2020, in Mississippi) were associated with a minimum of 20% decline in R_t_ in all three states, probably due to the early intervention.

**Figure 2:**
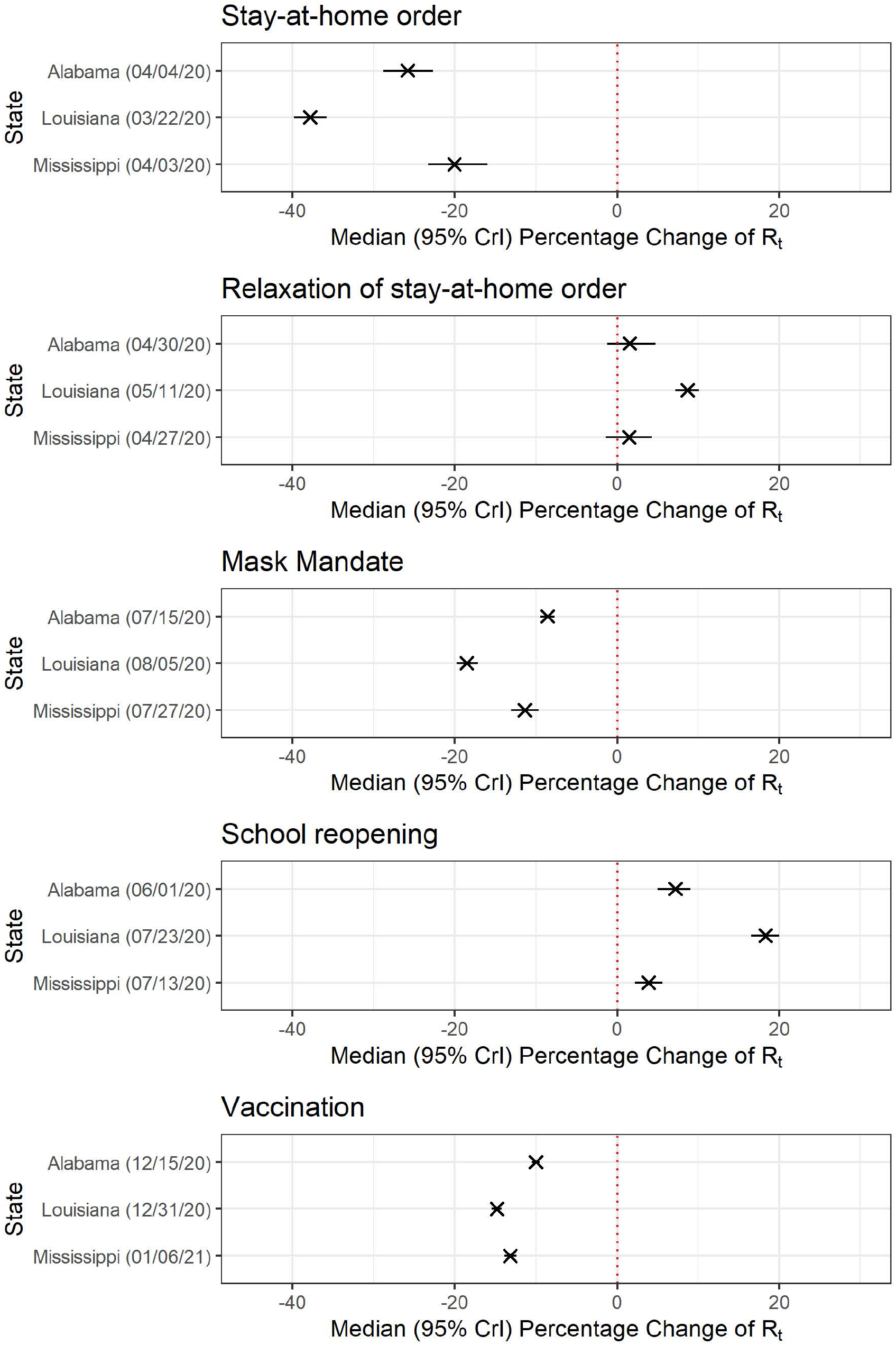
Median percentage change (95% credible intervals, CrI) of policy change R_t_ estimates for Alabama, Louisiana, and Mississippi grouped by social and public health interventions.

When the stay-at-home order was relaxed (represented as B), the R_t_ elevated in Louisiana by 8.69% (95% CrI: 7.19%, 10.09%), but the change was statistically insignificant in Alabama (1.56%, 95% CrI: -1.19%, 4.75%) and Mississippi (1.49%, 95% CrI: -1.41%, 4.27%). A possible reason was that the stay-at-home order was implemented in a rather relaxed manner, and its relaxation did not make a substantial behavioral change in human contact. On the contrary, when facemask mandates (represented as F) were enacted, there was a decline in the R_t_ by 8.55% (95% CrI: 7.68%, 9.41%), 18.51% (95% CrI: 1.75%, 17.16%), and 11.34% (95% CrI: 9.71%, 13.04%) in Alabama, Louisiana and Mississippi, respectively. Louisiana recorded the highest surge in transmission post-school reopening (represented by S) on July 23, 2020, with R_t_ increasing by 18.29% (95% CrI: 16.55%, 19.99%), followed by Alabama (7.19%, 95% CrI: 5.01%, 9.06%) and Mississippi (3.87%, 95% CrI: 2.22%, 5.59%). Our findings also suggested that the vaccination rollout (represented by V) against COVID-19 in all three states was associated with the median R_t_ values reduced to <1.

### Policy impacts at the county level

Following the enactment of the stay-at-home order in Alabama, Cullman County observed the highest percentage decrease in R_t_ by 53.55% (95% CrI: 21.79%, 71.73%). When the stay-at-home order was relaxed, Chambers and Cullman, observed an increase in R_t_ by 59.33% (95% CrI: 20.45%, 111.44%) and 67.89% (95% CrI: 14.07%, 161.52%) respectively. Interestingly, re-opening of schools and face mask mandates were not associated with a statistically significant change in R_t_ in all counties except Jefferson. Vaccination was associated with a dwindle in R_t_ in Cullman (−17.41%, 95% CrI: -21.14%, -13.58%) and Jefferson counties (−15.07%, 95% CrI: - 16.31%, -13.73%).

Louisiana had the lowest median R_t_ of 1.37 (95% CrI: 1.32, 1.42) among the three states before the implementation of the Stay-at-home order after the pandemic hit. After the stay-at-home order was enacted, Iberia observed the highest decline by 59.04% (95% CrI: 36.77%, 74.33%), then East Baton Rouge by 34.84% (95% CrI: 24.53%, 43.91%). The relaxation of the stay-at-home order was not associated with significant changes in R_t_ in any of the selected counties. In contrast, the face mask mandate was associated with an apparent decline in R_t_ in Iberia (−18.94%, 95% CrI: -27.07%, -10.40%) and East Baton Rouge (−17.26%, 95% CrI: -21.11%, - 12.86%). School reopening was associated with an increase in R_t_ in Iberia (18.84%, 95% CrI: 7.98%, 31.42%) and East Baton Rouge (16.31%, 95% CrI: 11.06%, 21.68%). Vaccination rollout was associated with a reduction in R_t_ by 9 to 14% in all three parishes.

In Mississippi, the stay-at-home order had the least impact in Hinds with a 19.69% (95% CrI: 4.88%, 33.08%) R_t_ reduction. The facemask mandate was not found to be associated with a change in R_t_ in Marshall and Leake counties. School reopening was followed by a surge in R_t_ in Hinds by 6.66% (95% CrI: 1.48%, 12.92%). Vaccination rollouts were associated with a statistically significant decline in R_t_ across all counties. Details of county-level policy impacts are presented in Tables S2, S3, and S4.

### Power-law Relationship between Cumulative Case Number and Population Size

Figure 3 and Table 1 present the results of the linear regression analysis between the log_10_-transformed per-capita cumulative incidence and the log_10_-transformed population size for all the counties in each of the three states. The negative slopes for Alabama on June 17 (−0.3229, 95% CI: -0.4964, -0.1495) and October 17, 2020 (−0.0820, 95% CI: -0.1404, -0.0236), suggested that in 2020, rural counties were experiencing a higher case burden than urban counties, whereas such disparity was not observed in the first half of 2021. The negative slopes for Louisiana and Mississippi at all four assessed dates suggested that low-population counties experienced a higher case burden throughout the study period.

**Table 1.** The slope (and 95% Confidence Intervals) of the linear regression line between log_10_-transformed per capita cumulative case number and log_10_-transformed population size, by state, Alabama, Louisiana, and Mississippi, on June 17, 2020, October 17, 2020, February 17, 2021, and May 17, 2021 (date of report).

| State | June 17, 2020 | October 17, 2020 | February 17, 2021 | May 17, 2021 |
| --- | --- | --- | --- | --- |
| Alabama | -0.3229<br>(-0.4964, -0.1495) | -0.0820<br>(-0.1404, -0.0236) | 0.0041<br>(-0.0418, 0.0499) | 0.0117<br>(-0.0318, 0.0553) |
| Louisiana | -0.0273<br>(-0.1812, 0.1266) | -0.0760<br>(-0.1332, -0.0189) | -0.0523<br>(-0.0901, -0.0146) | -0.0383<br>(-0.0742, -0.0024) |
| Mississippi | -0.2006<br>(-0.3837, -0.0175) | -0.1382<br>(-0.2013, -0.0749) | -0.0554<br>(-0.0945, -0.0164) | -0.0448<br>(-0.0808, -0.0089) |

**Figure 3:**
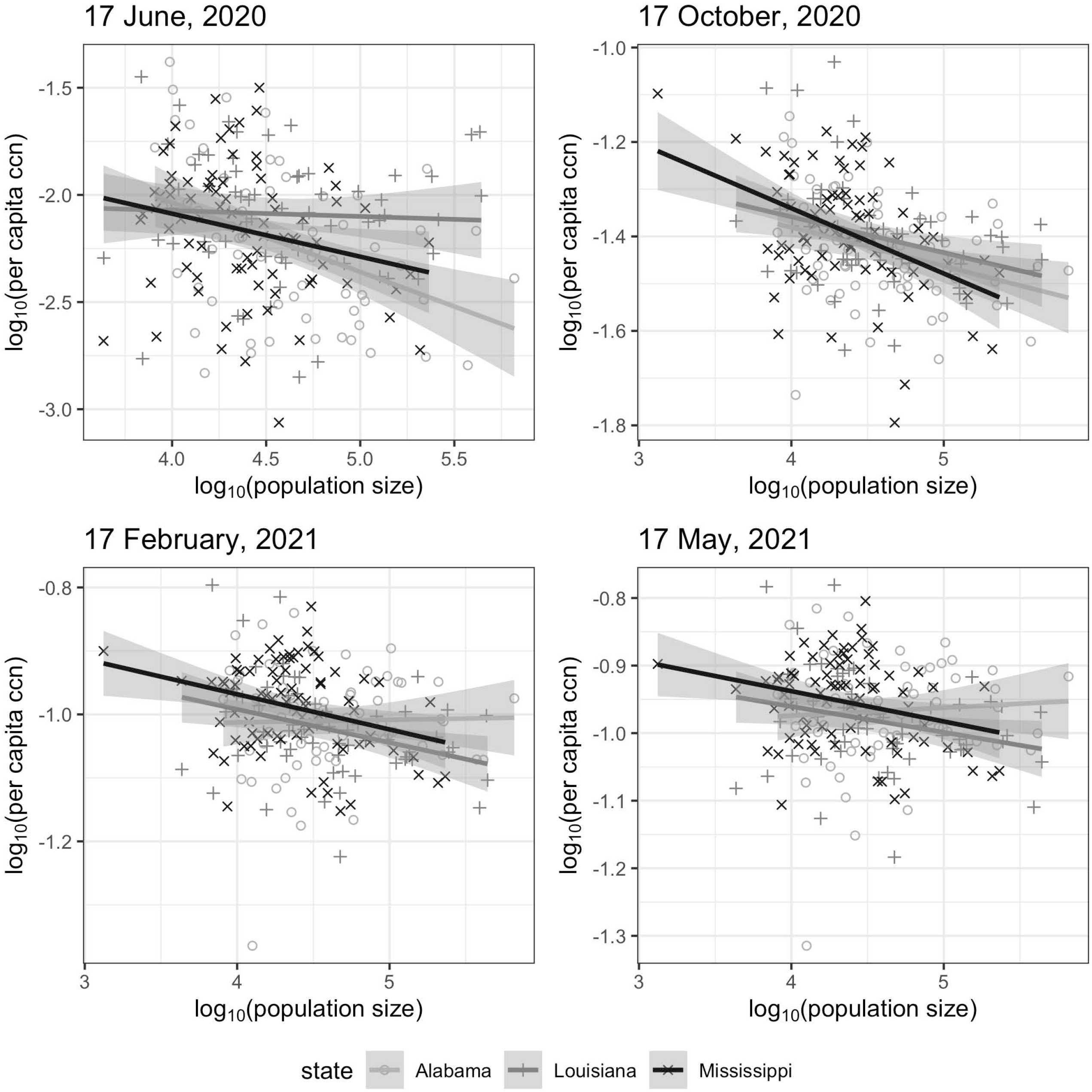
Linear regression plots of the relationship between log_10_-transformed per capita cumulative case number (ccn), and the log_10_-transformed population size for Alabama (light grey circle), Louisiana (grey cross), and Mississippi (black diagonal cross) by date of report on June 17, 2020, October 17, 2020, February 17, 2020 and May 17, 2020.

## Discussion

Overall, facemask mandates, stay-at-home orders, and vaccination rollout were the executive orders that were statistically significantly associated with decreased R_t_ values across all three states analyzed herein. School reopening was found to be associated with slightly increased transmission statewide, in Hinds county (100th percentile) in Mississippi, and in Iberia and East Baton Rouge parishes (75th and 100th percentile) in Louisiana. Meanwhile, the stay-at-home orders were associated with a decline in R_t_ in a majority of the selected counties. Our findings also suggest that counties with smaller population sizes were associated with a higher case burden throughout the study period for Louisiana and Mississippi and in the selected time points in 2020 for Alabama. Transmission appeared to be in decline after vaccines became available in the selected counties for each state except Chambers, Alabama. Counties with 100th percentile population size observed a significant decline in R_t_ following the stay-at-home order and face mask mandates.

Stay-at-home orders were issued in 43 of 50 states, when COVID-19 pandemic first hit the US in Spring 2020.^34^ These were primarily intended to reduce interpersonal contact and thus SARS-CoV-2 transmission, as demonstrated in prior studies.^35,36^ Other southern states like Georgia also experienced a significant decline in R_t_ to a value of <1 following the stay-at-home order.^30^ Another study to assess the effectiveness of stay-at-home orders in the US also found that such orders significantly reduced infection rates in Alabama, Louisiana, and Mississippi.^36^ In our study, Louisiana recorded the highest decline in R_t_ after this order probably due to the early implementation as confirmed in other studies.^37,38^ The relaxation of the order, therefore, led to an increase in R_t_ at the state level, and in Louisiana the increase was statistically significant. Underlying factors explaining the insignificant changes in transmission in Alabama and Mississippi should be explored in further studies.

School reopening has been reported by several studies to increase transmission of SARS-CoV-2.^39-41^ In our study, the Policy Change R_t_ increase after school reopening was statistically significant in Iberia and East Baton Rouge parishes, Louisiana (75^th^ and 100^th^ percentile population size) and Hinds, Mississippi (100^th^ percentile size). A mathematical modeling study on school reopening reported that it was associated with increased risk in urban regions.^42^ On the contrary, a study on COVID-19 in middle and high schools observed that counties with smaller population size in Florida were more likely to have an increased risk of transmission due to early reopening and a lack of mask mandates.^43^ Other studies in Michigan and Washington states also concluded the impact of school re-opening on SARS-CoV-2 depended on the community transmission potential.^44^ This is a probable explanation for the insignificant changes in Policy Change R_t_ in the five of the six selected counties with median and 75th percentile population sizes in our study.

In 2020, rural counties in Alabama, Louisiana and Mississippi had higher case burden than urban counties, similar to what was found in Georgia.^30^ To the contrary, the opposite was true in the non-Appalachian region of Kentucky. Meanwhile, rural-urban disparities was generally not observed in the Appalachian region of Kentucky and both Delta and non-Delta regions of Arkansas over much of 2020.^33^ In a study of 5 Western states,^41^ North Dakota was the only state where densely populated counties consistently had a higher per-capita cumulative incidence throughout 2020. Rural counties in Louisiana and Mississippi continued to experience a higher burden in the first half of 2021. This may be due to the low vaccination rates in these counties, poor compliance to public health measures, hospital closures, increased likelihood of unemployment, and delay in seeking care due to lack of insurance.^45,46^ This reiterates the need for public health outreach and the development of programs and policies to address the disparity.

### Limitations

First, the uncertainty associated with data accuracy and quality was a critical issue to consider. Data quality can be affected by testing policies of each state and the efficiency of the states’ case reporting systems. Second, the original dataset contained the dates of the case report and not the dates of infection or symptoms onset. Therefore, the epidemic curve was shifted backward by nine days to account for the incubation period (mean, 6 days) and delay to testing (median, 3 days). This method was considered “tolerable” by Gostic et al.^25^ We acknowledge that we did not use deconvolution,^47^ which was more computationally demanding, to approximate the date of infection. Third, this is an ecological study that identifies association but cannot demonstrate causality between public health policy and changes in R_t_. We were not able to conduct individual-level analysis due to the lack of demographic information of each case in aggregated data; hence, we could not investigate demographic risk factors for COVID-19 infection. Likewise, public health policies were implemented at a population level. Individuals’ compliance to policies might vary. Fourth, the comparison between three different states, in the same manner, may not be very accurate as test reporting could vary from state to state. Fifth, for county-level analysis, we did not choose county with population size below the median for comparison, because in our preliminary analysis (not shown), the low daily case count in counties with a small population would render the R_t_ estimates generated by the EpiEstim package very uncertain.

## Conclusions

Among all the policies implemented, the stay-at-home orders, face mask mandates, and vaccinations were associated with the most significant reductions in SARS-CoV-2 transmission in Alabama, Louisiana and Mississippi. The current study provides further evidence that state and county mandates and policy changes could have an impact on the trajectories of the pandemic in their jurisdictions. The rural-urban disparities in COVID-19 case burden reported here call for better evidence-based approaches in tailoring health promotion interventions and vaccination campaigns to rural residents and identifying the pertinent factors underlying the rural-urban disparities in the southern US.

## Data Availability

The cumulative incidence data for each county in Alabama, Louisiana, and Mississippi can be obtained from the New York Time GitHub data repository: https://github.com/nytimes/covid-19-data. All data produced in the present study are available upon reasonable request to the authors

## Supplementary Materials for

### Text S1: Additional details on Methods

#### Handling of negative incident case count

In instances of negative incident case count, i.e., when public health agencies made corrections, it was handled by correcting the incident case count of the previous day(s) and transforming the negative case count to zero. This is to ensure that the daily number of new cases matches the corrected cumulative case count of that day.

#### Cumulative case count and population size

The power-law relationship can be represented as:^1^

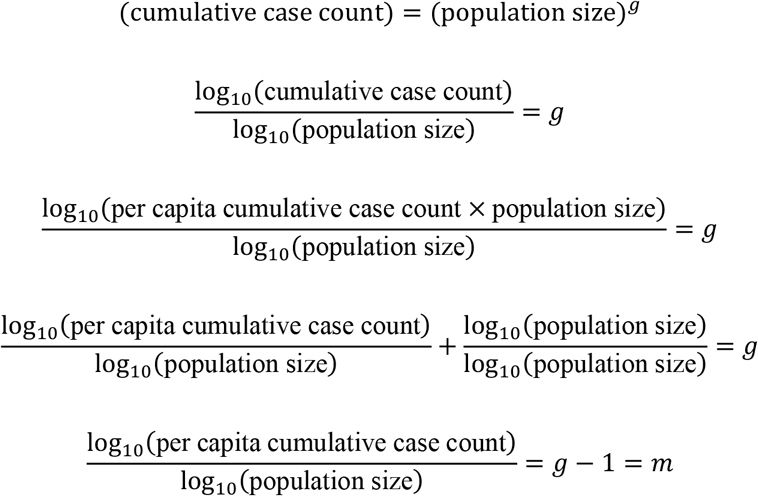

*m* is the slope of the regression line: *m*=1 means there is no heterogeneity of per capita cumulative incidence across the counties, *m*<1 means that counties with smaller population sizes have higher per capita incidence, while *m*>1 implies that the counties with larger population sizes have higher case burden.

### Text S2: Additional Results

#### Cumulative cases count by state and county

As of May 17, 2021, Alabama had recorded 540,267 confirmed cases, and for the selected counties, Jefferson recorded 79,879 cases, Cullman, 9,773 cases, and Chambers, 3,598 cases. Louisiana recorded a total of 465,946 cases, and at the parish level, East Baton Rouge had the highest case count (39,889), followed by Iberia (7,173) and Evangeline (3,625). In Mississippi, a total of 315,026 cases were reported (for the selected counties: Hinds: 28,049; Marshall: 28,093; Leake: 28,079) (Figures S1 and S2).

#### Spatial variation in confirmed COVID-19 cumulative case count and cumulative case count per 10,000 population

The cumulative case count and cumulative case count per 10,000 population across all counties on June 17 and October 17, 2020, and February 17 and May 17, 2021 (date of report) were presented in the maps (Figures S1 and S2).

As shown in Figure S1, the cumulative case count per 10,000 population indicated that Clarke (23,622 residents) and Hale (14,651 residents) counties in Alabama had the highest burden of COVID-19 with per capita cumulative incidence rates 1,488 and 1,529 per 10,000 population respectively, as of May 17, 2021. East Carroll (6,861 residents) and East Feliciana (19,539 residents) parishes in Louisiana recorded values of 1,647 and 1,657 per 10,000 population while Bolivar (30,985 residents) county in Mississippi had 1,569 per 10,000 population.^2^ These results support our conclusion that rural parishes/counties in Louisiana and Mississippi experienced a higher COVID-19 burden than urban parishes/counties.^3,4^ In Alabama, the relationship between per capita incidence and population size was rather homogeneous between rural and urban counties. For the two highest burden counties: Clarke is rural, whereas Hale is urban. This observation is supported by the finding from the regression analysis between log-transformed per capita cumulative case number and log-transformed population size as found in Table 1.

The maps of cumulative case count (Figure S2) show that as the pandemic progressed, Jefferson County (Birmingham) in Alabama had the highest cumulative case count of 79,879 as of May 17, 2021.

#### 7-day sliding window R_t_ estimates at the county level

In Alabama, the R_t_ for the county with the highest population size, Jefferson, decreased from 2.5 in March 2020 to below 1 in April, then fluctuated around one until it sustained a value of <1 between July and August 2020. The R_t_ values dropped to below one further in January, mid-February, March, and April 2020. The 75th and 50th percentile counties, Cullman, and Chambers, followed a similar trajectory. While there was a fluctuation in R_t_ throughout the study period, the values were lowest in March 2021 for Chambers and February 2021 in Cullman.

In Louisiana, all three selected parishes had R_t_ estimates of >3 in March, June, and November 2020. While the R_t_ was below 1 in East Baton Rouge (100th percentile population size) between July and October 2020, Evangeline (median population size) experienced another spike in mid-August 2020, and Iberia continued to fluctuate around 1.

In Mississippi, Hinds (100th percentile population size) recorded the lowest R_t_ (<1) in mid-February 2021 after a spike in June 2020. Across all three counties in Mississippi, the R_t_ surged (>1) between September and October 2020. Other than this, the R_t_ fluctuated around one throughout the study period for Marshall (75th percentile population size) and Leake (median population size) counties.

**Table S1.** The difference in Policy Change R_t_ as policies changed across Alabama, Louisiana, and Mississippi (State level).

| Alabama | Median $R_t$ (95% CrI) | Median $R_t$ difference and percentage changes comparing with previous policy interval (95% CrI) |
| --- | --- | --- |
| Before policy A 1 | 1.41 (1.36, 1.45) |  |
| A→B 1,2 | 1.04 (1.02, 1.07) | -25.81% (-28.78%, -22.69%) |
| B→S 2,3 | 1.06 (1.04, 1.08) | 1.56% (-1.19%, 4.75%) |
| S→F 3,4 | 1.14 (1.13, 1.15) | 7.19% (5.01%, 9.06%) |
| F→V 4,5 | 1.04 (1.04, 1.04) | -8.55% (-9.41%, -7.68%) |
| Beyond V 5+ | 0.92 (0.91, 0.92) | -9.97% (-10.47%, -9.47%) |

| Louisiana | Median $R_t$ (95% CrI) | Median $R_t$ difference and percentage changes comparing with previous policy interval (95% CrI) |
| --- | --- | --- |
| Before policy A 1 | 1.62 (1.57, 1.67) |  |
| A→B 1,2 | 1.00 (0.99, 1.02) | -37.82% (-39.80%, -35.76%) |
| B→F 2,3 | 1.09 (1.08, 1.10) | 8.68% (7.19%, 10.09%) |
| F→S 3,4 | 0.89 (0.88, 0.90) | -18.51% (-19.75%, -17.16%) |
| S→V 4,5 | 1.05 (1.05, 1.06) | 18.29% (16.55%, 19.99%) |
| Beyond V 5+ | 0.90 (0.89, 0.90) | -14.82% (-15.46%, -14.21%) |

| Mississippi | Median $R_t$ (95% CrI) | Median $R_t$ difference and percentage changes comparing with previous policy interval (95% CrI) |
| --- | --- | --- |
| Before policy A 1 | 1.37 (1.32, 1.42) |  |
| A→B 1,2 | 1.09 (1.07, 1.13) | -20.05% (-23.27%, -15.96%) |
| B→F 2,3 | 1.11 (1.10, 1.13) | 1.49% (-1.41%, 4.27%) |
| F→S 3,4 | 0.99 (0.97, 1.00) | -11.34% (-13.04%, -9.71%) |
| S→V 4,5 | 1.03 (1.02, 1.03) | 3.87% (2.22%, 5.59%) |
| Beyond V 5+ | 0.89 (0.88, 0.89) | -13.16% (-13.92%, -12.38%) |

CrI, credible intervals. Policy Change $R_t$ labels: A: Stay-at-home order, B: Safer-at-home order, S: School re-opening, F: Face mask mandate, V: Roll-out of vaccination began.

**Table S2.** The difference in Policy Change R_t_ as policies changed at county levels in selected counties in Alabama. These three counties were selected based on the 50^th^ percentile, 75^th^ percentile, and 100^th^ percentile population size.

| Chambers County (50<sup>th</sup>) | Median $R_t$ (95% CrI) | Median $R_t$ difference and percentage changes comparing with previous policy interval (95%CrI) |
| --- | --- | --- |
| Before policy A 1 | 1.43 (1.24, 1.64) |  |
| A→B 1,2 | 0.72 (0.59, 0.85) | -49.75% (-60.72%, -36.14%) |
| B→S 2,3 | 1.14 (0.92, 1.39) | 59.33% (20.45%, 111.44%) |
| S→F 3,4 | 1.03 (0.93, 1.14) | -9.599% (-27.60%, 13.9%) |
| F→V 4,5 | 1.03 (0.98, 1.09) | -0.36% (-11.05%, 11.07%) |
| Beyond V 5+ | 0.96 (0.91, 1.01) | -6.38% (-13.55%, 0.67%) |

| Cullman County (75<sup>th</sup>) | Median $R_t$ (95% CrI) | Median $R_t$ difference and percentage changes comparing with previous policy interval (95%CrI) |
| --- | --- | --- |
| Before policy A 1 | 1.57 (1.13, 2.12) |  |
| A→B 1,2 | 0.74 (0.495, 1.06) | -53.55% (-71.73%, -21.79%) |
| B→S 2,3 | 1.25 (1.06, 1.46) | 67.89% (14.07%, 161.52%) |
| S→F 3,4 | 1.13 (1.04, 1.21) | -9.69% (-24.95%, 9.06%) |
| F→V 4,5 | 1.06 (1.04, 1.09) | -5.45% (-12.32%, 2.06%) |
| Beyond V 5+ | 0.88 (0.85, 0.91) | -17.41% (-21.14, -13.58%) |

| Jefferson County (100<sup>th</sup>) | Median $R_t$ (95% CrI) | Median $R_t$ difference and percentage changes comparing with previous policy interval (95%CrI) |
| --- | --- | --- |
| Before policy A 1 | 1.23(1.13, 1.33) |  |
| A→B 1,2 | 0.99 (0.91, 1.08) | -19.17% (-28.22%, -9.031%) |
| B→S 2,3 | 1.08 (1.02, 1.15) | 8.75% (-1.70%, 21.12%) |
| S→F 3,4 | 1.14 (1.12, 1.17) | 5.71% (-0.82%, 12.79%) |
| F→V 4,5 | 1.06 (1.05, 1.07) | -7.29% (-9.69%, -5.04%) |
| Beyond V 5+ | 0.90 (0.89, 0.91) | -15.07% (-16.31%, -13.73%) |

CrI, credible intervals. Policy Change $R_t$ labels: A: Stay-at-home order, B: Safer-at-home order, S: School re-opening, F: Face mask mandate, V: Roll-out of vaccination began.

**Table S3.** The difference in Policy Change R_t_ as policies changed at parish levels in selected parishes in Louisiana. These three parishes were selected based on the 50^th^ percentile, 75^th^ percentile, and 100^th^ percentile population size.

| Evangeline Parish (50<sup>th</sup>) | Median $R_t$ (95% CrI) | Median $R_t$ difference and percentage changes comparing with previous policy interval (95% CrI) |
| --- | --- | --- |
| Before policy A 1 | 2.098 (0.89, 4.11) |  |
| A→B 1,2 | 0.96 (0.74, 1.22) | -55.06% (-77.24%, 15.55%) |
| B→F 2,3 | 1.13 (1.05, 1.22) | 17.44% (-8.98%, 52.32%) |
| F→S 3,4 | 1.02 (0.895, 1.15) | -9.92% (-22.57%, 5.02%) |
| S→V 4,5 | 1.01 (0.97, 1.06) | -0.59% (-13.49%, 14.02%) |
| Beyond V 5+ | 0.91 (0.85, 0.97) | -10.33% (-16.83%, -3.23%) |

| Iberia Parish (75<sup>th</sup>) | Median $R_t$ (95% CrI) | Median $R_t$ difference and percentage changes comparing with previous policy interval (95% CrI) |
| --- | --- | --- |
| Before policy A 1 | 2.57 (1.55, 3.95) |  |
| A→B 1,2 | 1.04 (0.94, 1.16) | -59.04% (-74.33%, -36.77%) |
| B→F 2,3 | 1.09 (1.04, 1.14) | 4.578% (-7.46, 17.44%) |
| F→S 3,4 | 0.88 (0.80, 0.97) | -18.94% (-27.07%, -10.40%) |
| S→V 4,5 | 1.05 (1.01, 1.09) | 18.84% (7.98%, 31.42%) |
| Beyond V 5+ | 0.91 (0.87, 0.95) | -13.80% (-18.78%, -8.63%) |

| East Baton Rouge Parish (100<sup>th</sup>) | Median $R_t$ (95% CrI) | Median $R_t$ difference and percentage changes comparing with previous policy interval (95% CrI) |
| --- | --- | --- |
| Before policy A 1 | 1.68 (1.45, 1.93) |  |
| A→B 1,2 | 1.09 (1.05, 1.13) | -34.84% (-43.91%, -24.53%) |
| B→F 2,3 | 1.07 (1.05, 1.098) | -1.36% (-5.90%, 2.66%) |
| F→S 3,4 | 0.89 (0.85, 0.93) | -17.26% (-21.11%, -12.86%) |
| S→V 4,5 | 1.03 (1.02, 1.05) | 16.31% (11.06%, 21.68%) |
| Beyond V 5+ | 0.93 (0.92, 0.95) | -9.63% (-11.73%, -7.47%) |

CrI, credible intervals. Policy Change $R_t$ labels: A: Stay-at-home order, B: Safer-at-home order, F: Face mask mandate, S: School re-opening, V: Roll-out of vaccination began.

**Table S4.** The difference in Policy Change R_t_ as policies changed at county levels in selected counties in Mississippi. These three counties were selected based on the 50^th^ percentile, 75^th^ percentile, and 100^th^ percentile population size.

| Leake County (50<sup>th</sup>) | Median $R_t$ (95%CrI) | Median $R_t$ difference and percentage changes comparing with previous policy interval (95%CrI) |
| --- | --- | --- |
| Before policy A 1 | 1.65 (1.10, 2.35) |  |
| A→B 1,2 | 1.20 (1.05, 1.36) | -26.95% (-50.09%, 10.43%) |
| B→F 2,3 | 0.96 (0.87, 1.05) | -20.14% (-31.31%, -6.27%) |
| F→S 3,4 | 0.91 (0.73, 1.13) | -5.42% (-24.79%, 19.52%) |
| S→V 4,5 | 1.03 (0.98, 1.09) | 12.79% (-9.30%, 43.07%) |
| Beyond V 5+ | 0.88 (0.79, 0.96) | -15.08% (-23.94%, -5.85%) |

| Marshall County (75<sup>th</sup>) | Median $R_t$ (95%CrI) | Median $R_t$ difference and percentage changes comparing with previous policy interval (95%CrI) |
| --- | --- | --- |
| Before policy A 1 | 1.28 (0.87, 1.79) |  |
| A→B 1,2 | 0.86 (0.53, 1.31) | -33.72% (-64.21%, 14.69%) |
| B→F 2,3 | 1.15 (1.03, 1.27) | 33.86% (-11.61%, 120.28%) |
| F→S 3,4 | 1.07 (0.92, 1.22) | -7.19% (-22.4%, 11.17%) |
| S→V 4,5 | 1.02 (0.99, 1.06) | -4.03% (-17.37%, 11.63%) |
| Beyond V 5+ | 0.92 (0.86, 0.97) | -10.90% (-16.93%, -4.35%) |

| Hinds County (100<sup>th</sup>) | Median $R_t$ (95%CrI) | Median $R_t$ difference and percentage changes comparing with previous policy interval (95%CrI) |
| --- | --- | --- |
| Before policy A 1 | 1.34 (1.17, 1.52) |  |
| A→B 1,2 | 1.07 (0.96, 1.19) | -19.69% (-33.08%, -4.88%) |
| B→F 2,3 | 1.13 (1.09, 1.17) | 5.46% (-5.35%, 17.77%) |
| F→S 3,4 | 0.95 (0.91, 1.00) | -15.68% (-20.83%, -10.28%) |
| S→V 4,5 | 1.02 (0.99, 1.04) | 6.66% (1.48%, 12.92%) |
| Beyond V 5+ | 0.90 (0.88, 0.93) | -11.3% (-14.34%, -8.16%) |

CrI, credible intervals. Policy Change $R_t$ labels: A: Stay-at-home order, B: Safer-at-home order, F: Face mask mandate, S: School re-opening, V: Roll-out of vaccination began.

## Supplementary Figure Legends

**Figure S1:**
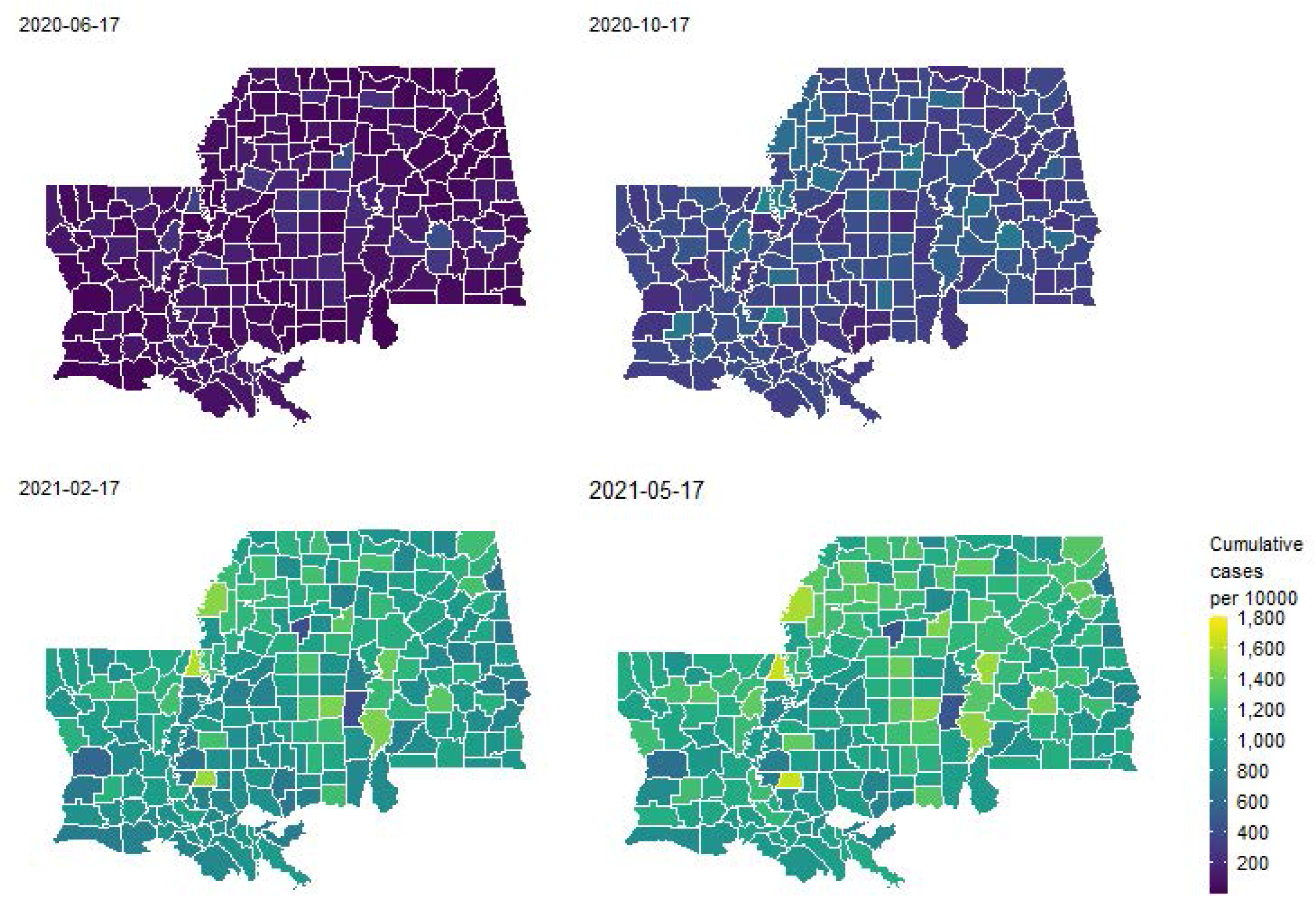
Maps of Alabama, Louisiana, and Mississippi: cumulative COVID-19 confirmed case count per 10,000 population by county: June 17, 2020, October 17, 2020, February 17, 2021, and May 17, 2021.

**Figure S2:**
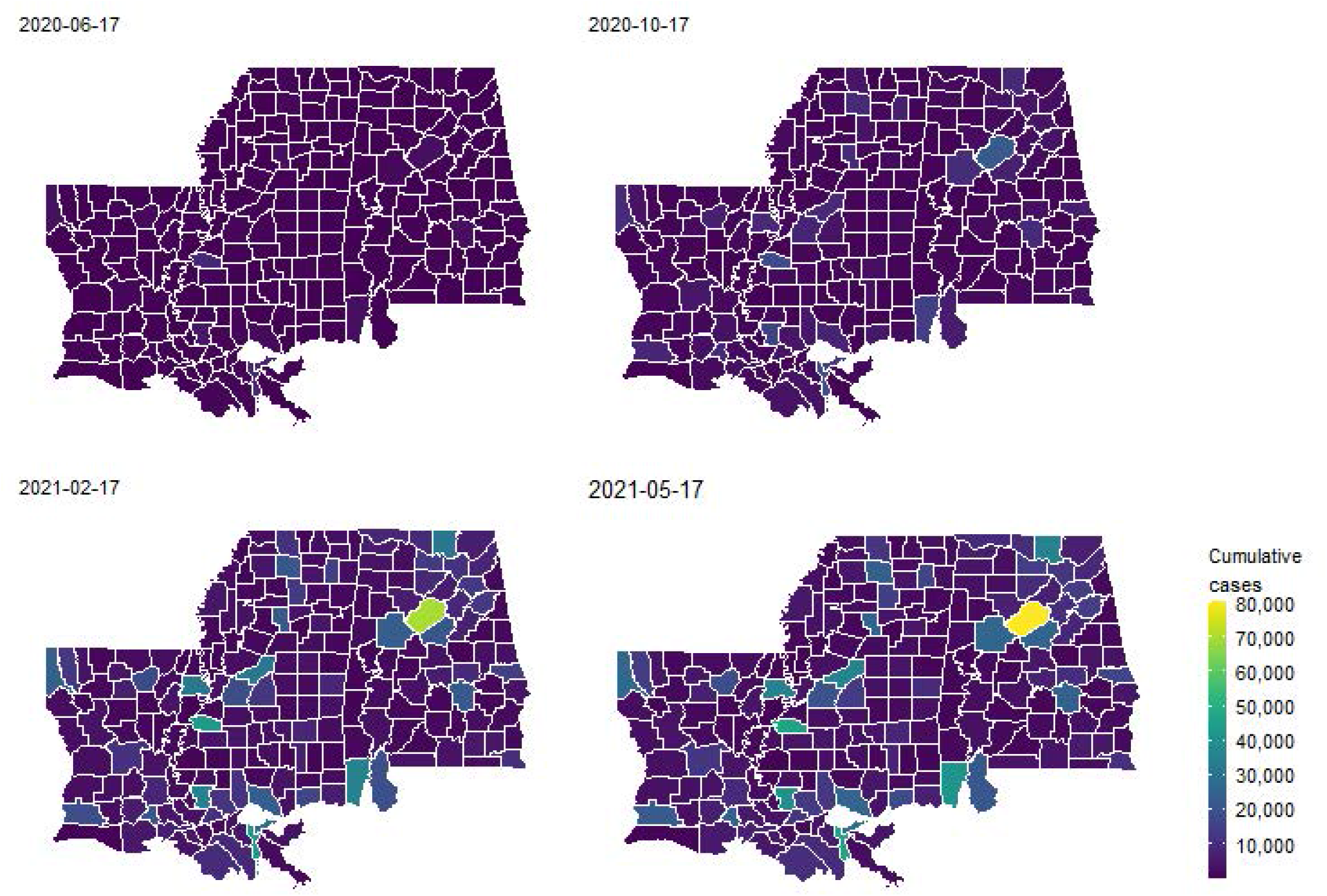
Maps of Alabama, Louisiana, and Mississippi: cumulative COVID-19 confirmed case count by county: June 17, 2020, October 17, 2020, February 17, 2021, and May 17, 2021.

**Figure S3:**
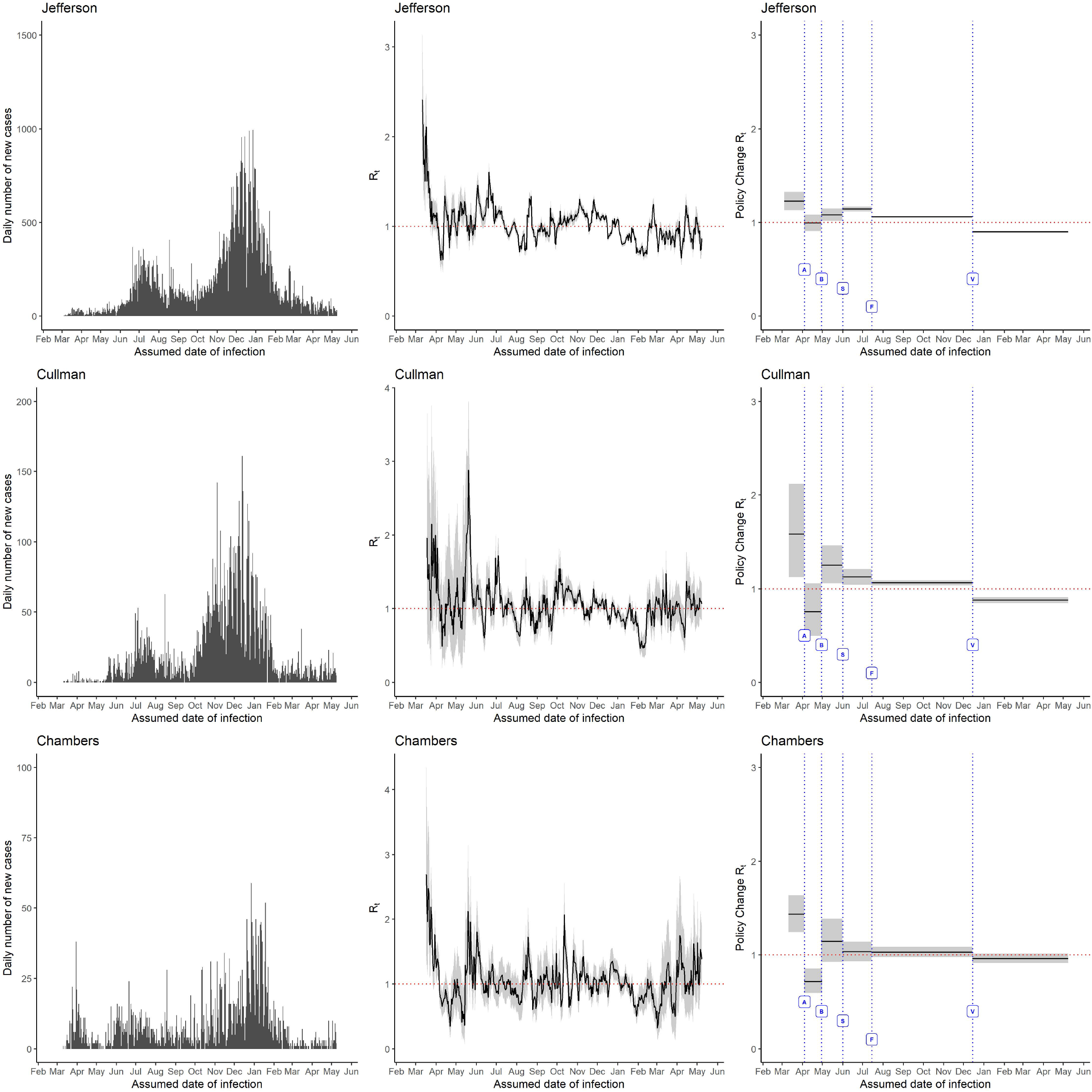
The daily number of new cases (left panel), 7-day sliding window R_t_ (middle panel), and non-overlapping window R_t_ for policy change (right panel) for counties with 100th percentile (Jefferson), 75th percentile (Cullman), and median (Chambers) population sizes in Alabama. The government policies represented by the alphabets in the figure are: A= Stay at home order directive, B= Shelter in place/safer at home, S= School re-opening, F= Face mask mandate, and V= Rollout of vaccination began.

**Figure S4:**
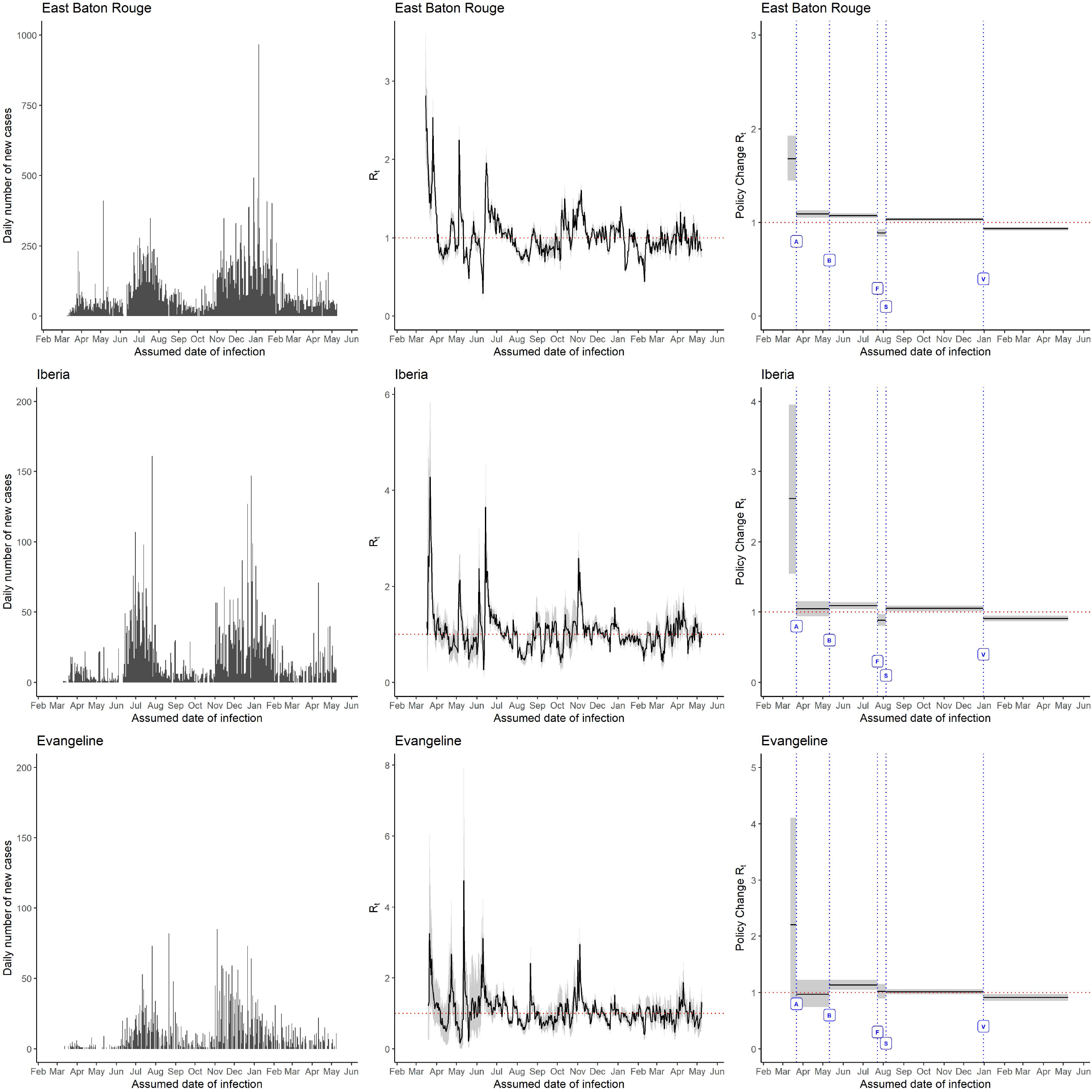
The daily number of new cases (left panel), 7-day sliding window R_t_ (middle panel), and non-overlapping window R_t_ for policy change (right panel) for counties with 100th percentile (East Baton Rouge), 75th percentile (Iberia) and 50th percentile (Evangeline) population sizes in Louisiana. The government policies represented by the alphabets in the figure are: A= Stay at home order directive, B= Shelter in place/safer at home, S= School re-opening, F= Face mask mandate, and V= Rollout of vaccination began.

**Figure S5:**
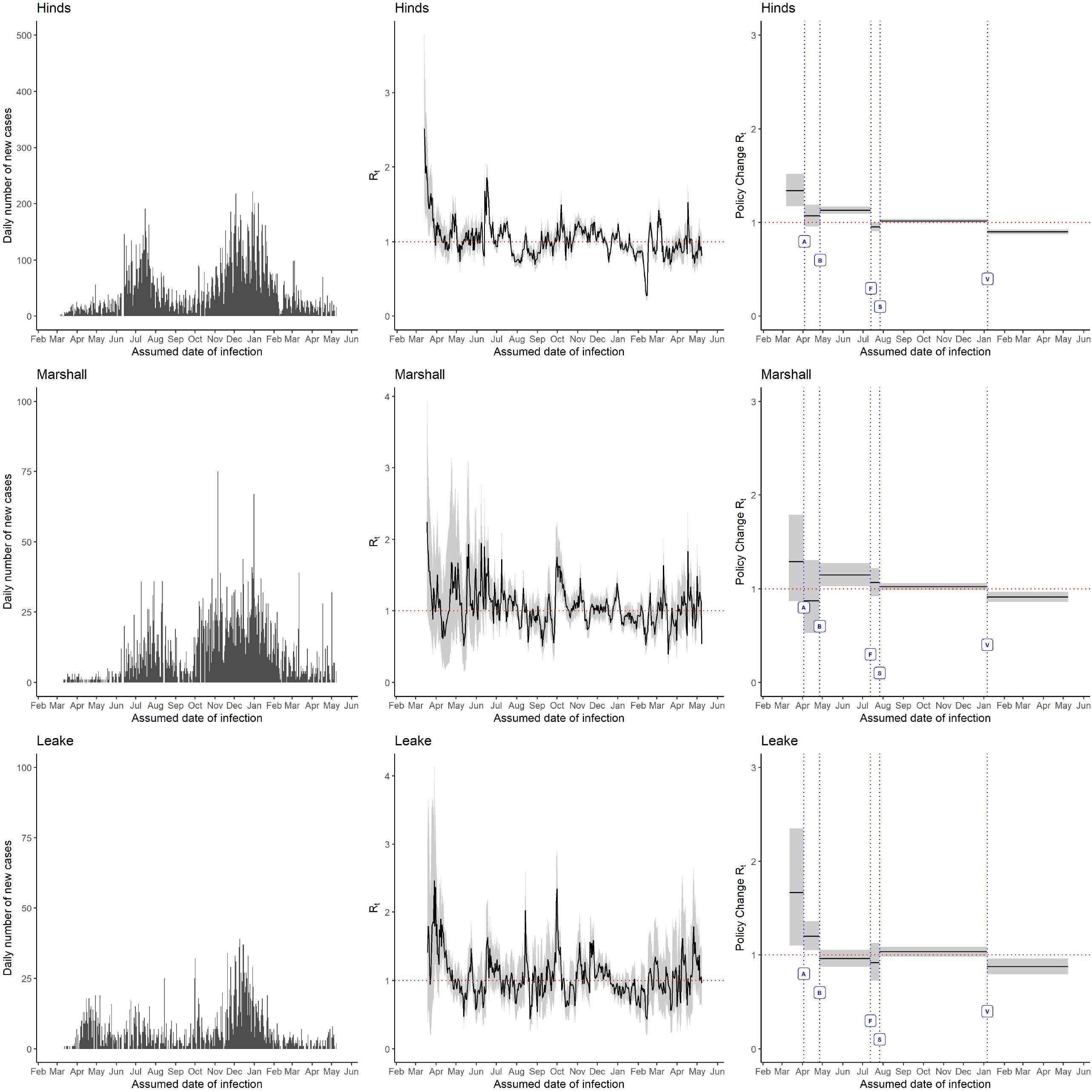
The daily number of new cases (left panel), 7-day sliding window R_t_ (middle panel), and non-overlapping window R_t_ for policy change (right panel) for counties with 100^th^ percentile (Hinds), 75th percentile (Marshall), and 50th percentile (Leake) population sizes in Mississippi. The government policies represented by the alphabets in the figure are: A= Stay at home order directive, B= Shelter in place/safer at home, S= School re-opening, F= Face mask mandate, and V= Rollout of vaccination began.

